# Reconsidering the case against risk prediction in self-harm: routinely collected health data distinguishes groups at higher and lower risk of adverse outcomes following paracetamol overdose

**DOI:** 10.64898/2026.07.15.26358127

**Authors:** J Oxley, L Schölin, G Brennan, A Anand, J Brett, M Eddleston, C Humphries

**Affiliations:** University of Edinburgh Centre for Population Health Sciences, Edinburgh, UK; University of Edinburgh Centre for Cardiovascular Science, Edinburgh, UK; NHS Lothian Department of Psychological Medicine, Edinburgh, UK; Drug Health Service, St Vincent’s Hospital Sydney, New South Wales, Australia; UNSW School of Clinical Medicine, New South Wales, Australia; University of Edinburgh Generative AI Laboratory, Edinburgh, UK

## Abstract

**Background:** UK clinical guidance recommends that structured risk prediction tools and risk stratification should not be used in self-harm, to predict suicide or determine who is offered treatment. Underpinning this position is the premise that routinely collected health data contain no useful predictive signal, which has received little direct scrutiny.

**Objective:** To test whether routinely collected electronic health record data can distinguish groups at higher and lower risk of severe outcomes following paracetamol overdose.

**Methods:** We analysed 4,095 adults presenting to NHS Lothian emergency departments with paracetamol overdose (2017-2023). Elastic-net logistic regression was fitted to 37 routinely collected electronic health record features to predict a composite of death or mental health inpatient admission at 0-7, 8-30 and 31-365 days following attendance, evaluated on a held-out 20% test set with bootstrapping.

**Findings:** Events occurred in 5.5% of patients at 0-7 days, 2.0% at 8-30 days and 7.9% at 31-365 days, dominated by mental health admission. Bootstrap AUROC 95% confidence intervals lay above 0.5 in every window (0.65-0.82, 0.63-0.90, 0.71-0.85): models ranked patients better than chance. Calibration slopes (1.04, 1.14, 1.07) were close to one. Ranking drew primarily on mental health-related features.

**Conclusions:** Routinely collected health data carried predictive signal for severe outcomes after paracetamol overdose, although discrimination fell short of what is needed for individual-level clinical use.

**Clinical implications:** These models are not proposed for clinical deployment; however, treating risk prediction as a settled question will redirect research efforts, potentially excluding this patient population from machine learning advances driving improvements in care in other medical specialties.

**Summary box:** *What is already known on this topic:* NICE NG225 (2022), NHS England’s Staying Safe from Suicide framework (2025) and NCISH guidance (2024) recommend against structured risk prediction in self-harm, both for predicting suicide or repetition and for allocating treatment. An increasingly common reading of the underpinning literature is that routinely collected health data contain no useful predictive signal in this population. Evaluations of this claim in large-scale UK linked healthcare datasets remain scarce.

*What this study adds:* In a whole-population cohort of adults attending emergency departments with paracetamol overdose, models using 37 structured electronic health record features separated groups at higher and lower risk of all-cause mortality or mental health admission across all three time horizons, with calibration slopes close to one. Discriminative signal was carried primarily by mental health-related features rather than demographics. Useful predictive signal is therefore recoverable from routinely collected UK healthcare data in this population.

*How this study might affect research, practice or policy:* These findings are not evidence that suicide can be predicted at the individual level, and the models are not proposed for clinical use. They do, however, indicate that the empirical question underlying current guidance remains open, and should stay subject to scientific enquiry and to revision if sufficiently robust models emerge. Future research should expand the feature space beyond structured records, particularly into clinical free text, where the information on which psychosocial assessment relies already resides.

## Introduction

People who present to hospital after self-poisoning are at elevated risk of mortality and rehospitalisation in the year that follows presentation. In the United Kingdom, paracetamol is the drug most frequently implicated in deliberate self-poisoning, and the rates of paracetamol use as a method of self-harm are likely the highest of any nation^1–4^. The clinical pathway that follows presentations of paracetamol overdose combines medical management with a psychosocial assessment intended to identify patients requiring further mental health input.

Poor outcomes following self-harm assessment pathways are common. A systematic review of hospital-presenting self-harm reports a one-year repetition rate of approximately 17% and a one-year suicide rate of approximately 1.6%^5^. The National Confidential Inquiry into Suicide and Safety in Mental Health (NCISH) annual report has further documented that most people who die by suicide were assessed as at low or no immediate risk at their last contact with mental health services^6^. Even with current NICE-recommended pathways, a substantial fraction of patients deteriorate or die in the year following self-harm presentation^5,6^. The potential for improvement in the identification of patients at risk of poor outcome following a presentation of self-harm is therefore considerable.

Over the past four years, English guidance on structured risk prediction in self-harm has converged on a restrictive position. NICE guideline NG225 (2022) recommends explicitly against the use of risk assessment tools and scales to predict suicide or repetition of self-harm, and against the use of low, medium or high risk stratification for the same purposes; parallel recommendations in the same guideline direct that neither tools nor stratification be used to determine who should be offered treatment or who should be discharged^7^. NCISH advises that such instruments “should not be used to predict suicide risk or to allocate treatment”^8^. In 2025, the NHS England Staying Safe from Suicide framework moved beyond the NICE position to declare risk stratification, as a category, “not acceptable”^9^. The Royal College of Psychiatrists, the Healthcare Safety Investigation Branch and coronial prevention of future deaths reports have converged on the same position^10^. The recommendations therefore extend beyond suicide prediction, covering the use of risk stratification for any purpose, including identification of patients at risk of severe outcomes.

NICE guidance retains psychosocial assessment by mental health professionals as the recommended care pathway following self-harm^7^. The recommendation against prediction applies only to structured risk assessment tools, not to the clinical assessment pathway itself: structured, formulation-based approaches to understanding an individual’s risk, needs and circumstances remain central to recommended practice. However, as data insights become more accessible via large-scale data linkage, and advances in machine learning improve complex pattern recognition, it is conceivable that risk prediction tools have the potential to become valuable components of clinical decision-making, supporting consistent application of evidence-based risk factors and helping to ensure equitable standards of care across healthcare settings. Current health policies risk directing scientific enquiry in this sphere away from emerging data science approaches, although investigation of predictive models and their clinical utility is distinct from advocating their immediate adoption into routine practice.

Current guidelines draw on several strands of evidence. A national audit found risk assessment instruments in NHS use to be short, locally produced and inconsistently applied; seminal prospective work showed individual-level prediction of suicide from single risk factors generates unmanageable false positive rates, and a 50-year meta-analysis found individual risk factors to be weak predictors^11–13^. The contemporary modelling literature is larger and more contested: meta-analyses of machine learning models report widely varying discrimination, systematic appraisal finds most published models at high risk of bias and very few externally validated, and methodologists have argued that several common criticisms of prediction models rest on outdated, classification-based thinking^14–17^. The question of what predictive information routinely collected data contain is therefore live rather than settled.

This paper asks whether routinely collected electronic health record (EHR) data available at the point of emergency department presentation contain predictive information for adverse outcomes over the subsequent year, in patients presenting after paracetamol overdose. Our objectives were to report the occurrence of a composite outcome of all-cause mortality or mental health inpatient admission in three follow-up windows after presentation (0-7, 8-30 and 31-365 day prediction horizons), and to quantify whether the discrimination and calibration of models built from routine data in each window were statistically superior to chance. The study tests the claim that no useful signal is recoverable from routine data. Whether suicide itself can be predicted at the individual level is a separate question that can only be answered if this line of enquiry remains open. Rather than an attempt to build or endorse a deployable tool, this study is a deliberate test of a single concept: whether any useful predictive signal is recoverable from routine data.

## Methods

### Study design and population

We conducted a population-based study using linked routinely collected electronic health records covering all NHS-provided care within NHS Lothian, UK (population approximately 900,000). Episode-level data from NHS emergency department, inpatient and outpatient encounters, community prescribing records, GP records and national mortality data are linked using a single patient identifier. Every feature considered for the model was conceivably recoverable from routine data to be made available to the treating clinician at the point of contact.

The eligible population comprised adults (≥16 years) attending any NHS Lothian emergency department between 1 November 2017 and 31 May 2023 with a discharge diagnosis coded as “paracetamol overdose”. Each patient contributed a single index attendance, defined as their first such presentation in the study period; a 12-month lookback for candidate predictor derivation and a 12-month follow-up for outcome ascertainment were applied around each index attendance.

### Outcome

The primary outcome was a composite of all-cause mortality or any mental health inpatient admission in one of three follow-up windows: 0-7, 8-30 and 31-365 days after the index ED attendance, reflecting established time periods for outcome studies following ED attendance^18^. Each window was treated as a separate prediction problem with the outcome defined as event occurrence specifically within that window. Patients with a mental health admission or death in an earlier window remained eligible in later windows; this decision reflects how a clinician would apply prediction prospectively, without knowledge of future events. Per-horizon event rates therefore represent the marginal probability of an event in the specified window and are not cumulative incidence. Follow-up windows were anchored to the date of the index ED attendance rather than to hospital discharge. A composite outcome was used as admission to a mental health unit captures clinically high-risk deterioration short of death.

We constructed three separate models rather than a single time-to-event model because the predictors plausibly operate over different mechanisms at each horizon. Modelling each window separately preserves the full at-risk denominator at every horizon and avoids imposing a single proportional-hazards structure across mechanisms that plausibly differ.

### Candidate predictors

Thirty-seven candidate predictors were chosen by clinician feature selection: each was a routinely recorded measure which was conceivably informative for subsequent presentations: demographics (age, sex, Scottish Index of Multiple Deprivation [SIMD] quintile, SIMD missingness indicator), prior outpatient activity (any attendance flag, attended and did-not-attend status, six specialty flags including general, geriatric, child and adolescent, forensic and learning disability psychiatry, and clinical psychology), GP-recorded mental health and substance use diagnoses encoded as a phenotype list (alcohol problems, alcoholic liver disease, anxiety, depression, schizophrenia, bipolar disorder, personality disorder, OCD, autism, ADHD, eating disorders, dementia, delirium, intellectual disability, other psychiatric and substance misuse codes), and community prescribing (binary flags for prescriptions of antidepressants, hypnotics/anxiolytics, antiepileptics, antipsychotic-class drugs, substance dependence medication, CNS stimulants for ADHD, and analgesics; plus a continuous count of unique Virtual Therapeutic Moieties as a polypharmacy index). Absence of activity in the lookback period was treated as non-exposure rather than missing data, reflecting how the records are generated. Predictors were specified a priori on clinical grounds rather than by data-driven screening, so the candidate set itself introduces no selection instability; final selection within each training fold was performed by the elastic-net penalty.

Missingness was minimal. Age, sex and date of death were fully populated. SIMD was missing for 96 of 4,095 patients (2.3%); we retained a missingness flag predictor in case these data were missing-not-at-random due to housing status, and missing values were imputed as the median quintile within each cross-validation fold.

### Modelling

Three elastic-net logistic regression models were fitted, one per outcome window. The elastic-net penalty combines LASSO (which drives weak coefficients exactly to zero, performing feature selection) and ridge (which shrinks correlated coefficients toward each other, stabilising selection)^19^. The mixing parameter α and overall penalty λ were chosen by nested 5-fold cross-validation over a 10-point α grid (0.1-1.0), maximising AUROC. Balanced weighting was used, with subsequent Platt scaling for post-hoc probability calibration; the calibration model was fitted on out-of-fold predictions from the training set only, so that both the elastic-net and the recalibration model were evaluated on data they had not seen^20^. Balanced weighting compensates for the low event fraction during fitting; because it distorts the output probability scale, Platt scaling was applied afterwards, and all reported calibration metrics derive from these recalibrated probabilities.

The dataset was split into 80% training (n=3,275) and 20% held-out test (n=820) by stratified random sampling on a 365-day composite outcome. The test set was untouched during model selection, hyperparameter tuning, imputation and recalibration.

### Uncertainty quantification

Two performance properties of a probabilistic model were reported throughout. Discrimination was the model’s ability to rank patients who go on to experience the outcome ahead of those who do not, summarised by the area under the receiver-operating-characteristic curve (AUROC): 0.5 indicates ranking indistinguishable from chance and 1.0 indicates perfect ranking. Calibration is whether assigned probabilities match observed frequencies: a calibration intercept of zero indicates no systematic over- or under-estimation and a slope of one indicates that probabilities are credible.

A 1,000-iteration bootstrap was used to derive 95% confidence intervals around the AUROC, calibration intercept and slope and around the proportion of iterations in which each candidate feature was retained by the elastic-net penalty.

All analysis was performed in R (v4.5.3) using rsample (v1.1.1) for splitting, glmnet (v4.1-7) for model fitting, and stats (v4.5.3) for the calibration models. A fixed random seed was used throughout to ensure reproducibility of all splits, bootstrap resamples and cross-validation folds. The manuscript follows the TRIPOD+AI reporting guideline, with the completed checklist provided as a separate submission document.

### Ethical approval

Ethical approval was granted by DataLoch (DL-2024-044). All published outputs comply with NHS Scotland statistical disclosure control; values with counts <10 are suppressed.

## Results

### Cohort

6,763 attendances were coded for a paracetamol overdose discharge diagnosis, corresponding to 4,095 unique adult patients meeting inclusion criteria (Table 1). The cohort was 61.2% female, with a median age of 33 years (IQR 24-49). 51.7% of patients fell in SIMD quintiles 1 or 2, consistent with a social deprivation gradient reported in earlier Scottish data^21^. In the 12-month lookback, 50.2% of patients received an antidepressant prescription, 23.5% a hypnotic or anxiolytic, 13.3% an antipsychotic-class prescription and 6.7% a prescription for substance dependence. 44.5% had at least one prior outpatient appointment and 41.2% had a prior general psychiatry outpatient contact. Stratified random sampling produced training (n=3,275) and held-out test (n=820) sets balanced on outcome prevalence.

**Table 1.** Baseline characteristics of the overall cohort (n=4,095). All values are reproduced directly from disclosure-controlled summary outputs; cells suppressed under NHS Scotland Statistical Disclosure Control are shown as “< 10”.

| Characteristic | Overall cohort (n=4,095) |
| --- | --- |
| Female sex, n (%) | 2,506 (61.2) |
| Male sex, n (%) | 1,589 (38.8) |
| Age, years; median (Q1-Q3) | 33 (24-49) |
| SIMD quintile 1 (most deprived), n (%) | 909 (22.2) |
| SIMD quintile 2, n (%) | 1,207 (29.5) |
| SIMD quintile 3, n (%) | 709 (17.3) |
| SIMD quintile 4, n (%) | 573 (14.0) |
| SIMD quintile 5 (least deprived), n (%) | 601 (14.7) |
| SIMD missing, n (%) | 96 (2.3) |
| Unique VTM medications prescribed in lookback; median (Q1-Q3) | 1 (0-2) |
| Any prior outpatient attendance, n (%) | 1,821 (44.5) |
| General psychiatry outpatient contact, n (%) | 1,686 (41.2) |
| Antidepressant prescription, n (%) | 2,054 (50.2) |
| Hypnotic / anxiolytic prescription, n (%) | 963 (23.5) |
| Antipsychotic-related prescription, n (%) | 546 (13.3) |
| Antiepileptic prescription, n (%) | 451 (11.0) |
| Substance dependence prescription, n (%) | 274 (6.7) |
| GP-coded depression, n (%) | 336 (8.2) |
| GP-coded anxiety, n (%) | 251 (6.1) |
| GP-coded alcohol problems, n (%) | 191 (4.7) |
| GP-coded other psychiatric / substance misuse, n (%) | 186 (4.5) |
| GP-coded personality disorder, n (%) | 125 (3.1) |
| GP-coded schizophrenia, n (%) | 25 (0.6) |
| GP-coded autism, n (%) | 20 (0.5) |
| GP-coded bipolar disorder, n (%) | 18 (0.4) |
| GP-coded eating disorder, n (%) | 16 (0.4) |
| GP-coded dementia, n (%) | 15 (0.4) |
| GP-coded ADHD, n (%) | 12 (0.3) |
| GP-coded alcoholic liver disease, n (%) | < 10 |
| GP-coded intellectual disability, n (%) | < 10 |
| GP-coded OCD, n (%) | < 10 |
| GP-coded non-substance delirium, n (%) | < 10 |

Composite outcome events occurred in 224 patients (5.5%) in the 0-7 day window, 82 (2.0%) in the 8-30 day window and 325 (7.9%) in the 31-365 day window. Mental health admission dominated the composite in every window; per-component counts are withheld under statistical disclosure control.

### Discrimination and calibration

Held-out test-set AUROCs and corresponding bootstrap-median AUROCs over 1,000 resamples of the training data are shown in Figure 1. The bootstrap 95% confidence interval lay entirely above 0.5 at every horizon, indicating discrimination above chance under repeated resampling. The 8-30 day window has the widest interval, reflecting the low event count at that horizon rather than a substantive performance difference.

**Figure 1.**
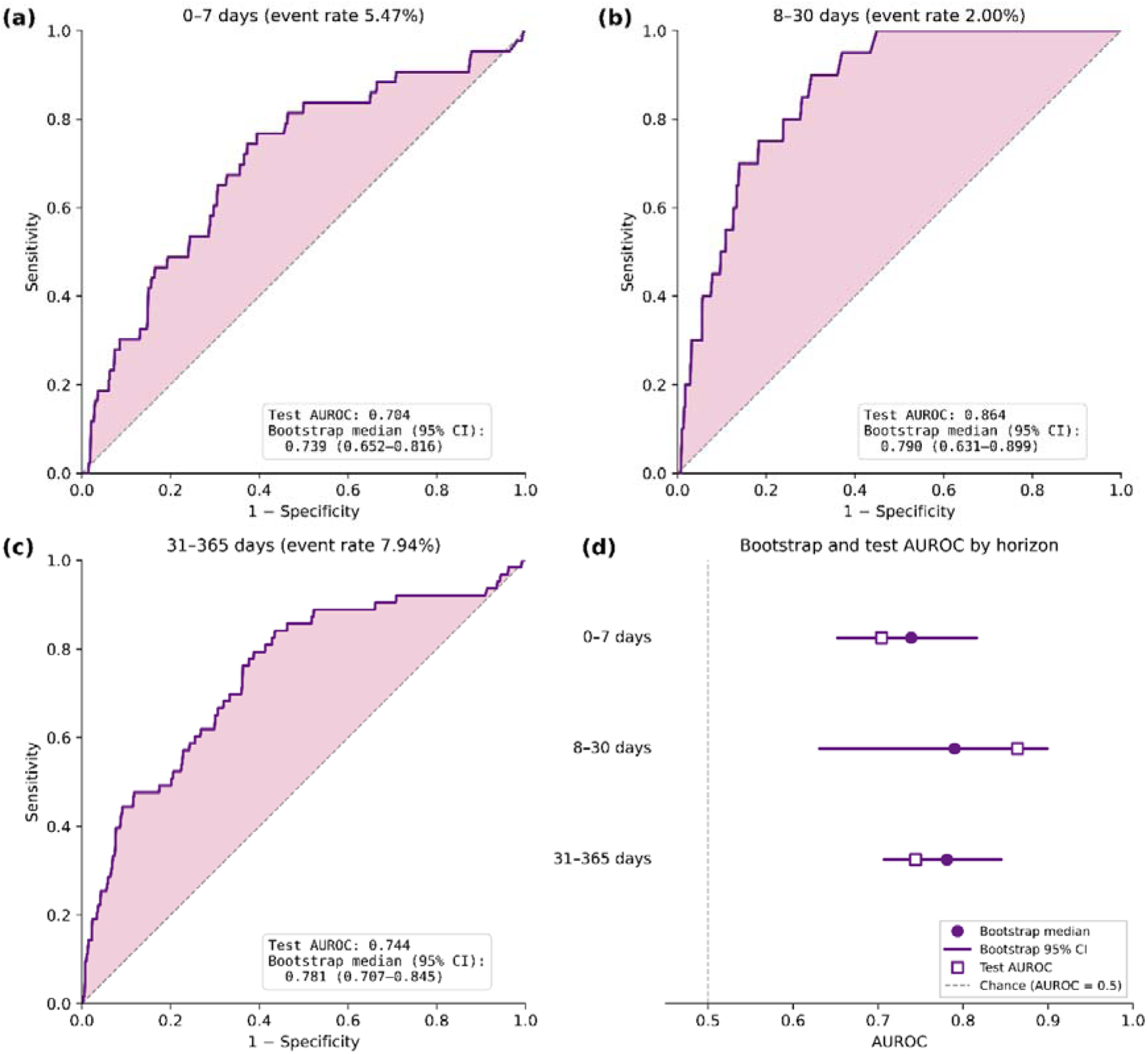
Discrimination of the three elastic-net composite outcome models on the 20% held-out test set. The dark curve is the held-out ROC; with the area between the ROC curve and the chance diagonal shaded to illustrate the separation from chance. Each panel reports test set AUROC, and the training set median AUROC with bootstrap 95% confidence intervals. (a) 0-7 day window, (b) 8-30 day window, (c) 31-365 day window, (d) bootstrap-median AUROC with 95% confidence interval and held-out test AUROC, by follow-up window.

Bootstrap-median calibration slopes were 1.04, 1.14 and 1.07, all close to one. Bootstrap-median calibration intercepts were +0.10, +0.47 and +0.14; the 95% confidence interval for the intercept included zero in each model, with widths of −1.30 to +2.01 at 0-7 days, −2.34 to +8.19 at 8-30 days, and −0.77 to +1.21 at 31-365 days. The wide interval in the 8-30 day window again reflects the low event rate at that horizon. Held-out raw calibration intercepts of −0.42, +0.67 and −0.19 lay within the corresponding bootstrap intervals; Platt-scaled Brier scores were 0.050, 0.023 and 0.066^22^.

**Figure 2.**
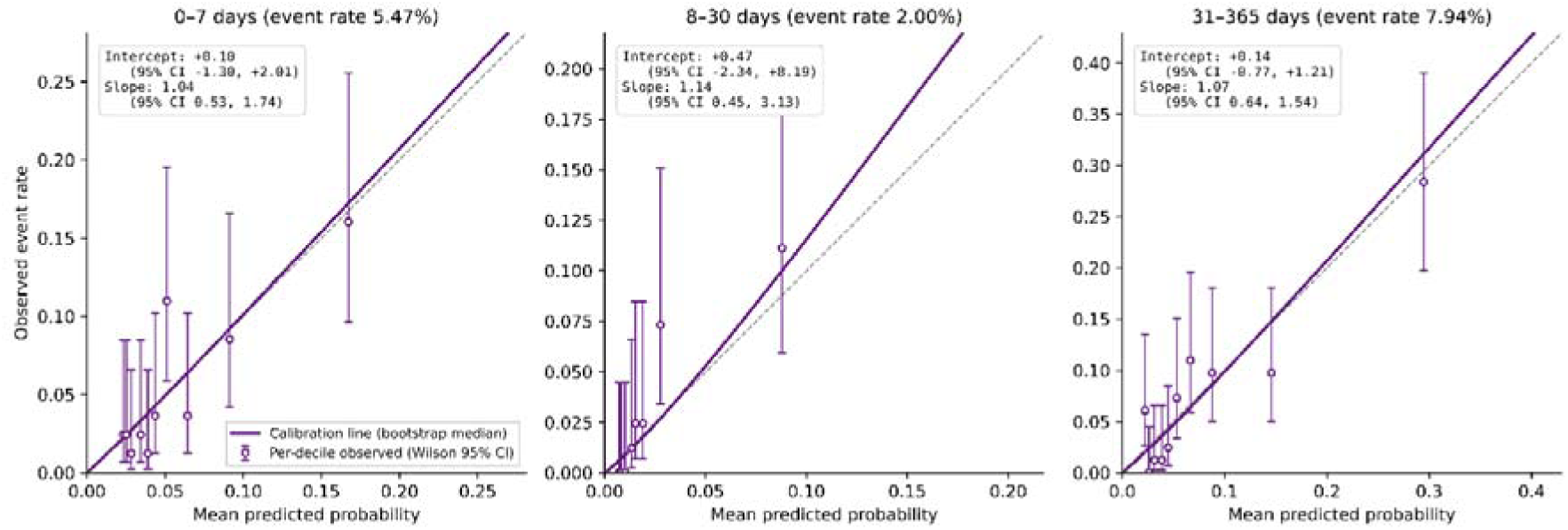
Calibration on held-out test data. Open circles are the per-decile observed event rates with Wilson 95% binomial confidence intervals (vertical bars), giving the per-decile sampling uncertainty. The dark purple line is the bootstrap-median logistic calibration curve y(x) = invlogit(intercept + slope · logit(x)), evaluated at the bootstrap-median intercept and slope as a smooth-fit summary. The dashed line indicates perfect calibration. Bootstrap median and 95% CI for intercept and slope are reproduced in each panel.

### Feature retention

Different combinations of features were retained at different horizons (Figure 3). The 0-7 day model retained six features dominated by markers of severe mental illness and polypharmacy. The 8-30 day model, with the smallest event count, was the most heavily penalised (alpha=1.0, pure LASSO) and retained only three features: geriatric psychiatry outpatient contact, alcoholic liver disease and polypharmacy. The 31-365 day model retained eight features blending prior service contact, psychiatric and substance dependence prescribing, age, polypharmacy and total GP-diagnostic burden.

**Figure 3.**
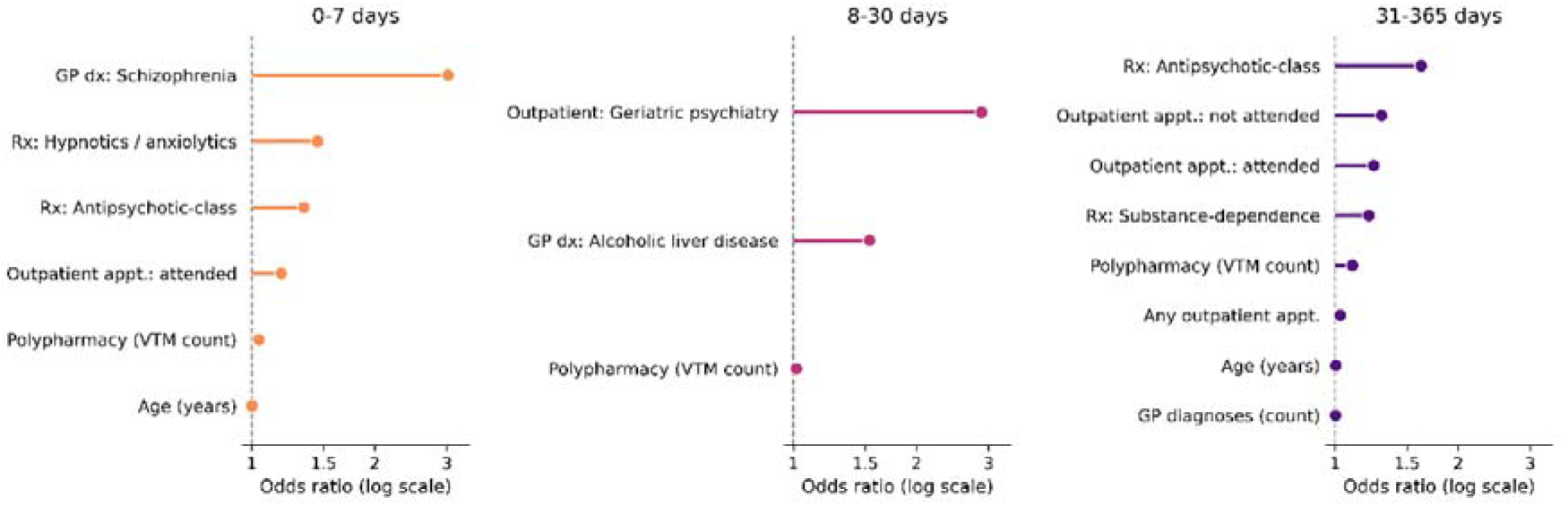
Retained features (non-zero standardised coefficients) from the final elastic-net models, by follow-up window. All non-zero coefficients are positive: each retained feature is associated with increased odds of the composite outcome. Specific features should be interpreted as illustrative of the feature classes the model draws on (severe-MH service contact, prior MH-specialty activity, polypharmacy/multimorbidity, substance misuse) rather than as a definitive ranking. Bootstrap retention frequencies are tabulated in the supplement. Axis is log-odds, dashed line at odds ratio of 1 signifies no effect.

Bootstrap retention frequencies (Figure 4; complete tables in the supplement) characterise which candidate predictors contributed reliably across resamples of the training data. Age was retained in nearly every resample at the 0-7 day and 31-365 day horizons (99.7% and 99.9% respectively) and in 26.7% of resamples at 8-30 days. Sex was retained in fewer than half of resamples at every horizon (45.4%, 29.7% and 39.3%); area-level deprivation (SIMD quintile) was retained in fewer than half at the 0-7 and 8-30 day horizons (41.7% and 39.8%) and in 56.7% of resamples at 31-365 days. Panel B of Figure 4 places these retention frequencies alongside per-predictor effect sizes. The disparity is informative: age, although retained at very high frequency, carried a median bootstrap coefficient near zero at every horizon (≤0.02), whereas several mental health-related features carried substantially larger effects (median bootstrap coefficient 0.4 to 2.2 for antipsychotic-class prescribing, GP-coded schizophrenia, hypnotic and anxiolytic prescribing, and alcoholic liver disease in at least one horizon). High retention of age therefore does not imply that age was the predominant source of discriminative signal; the held-out discrimination drew on the combination of demographic and mental health-related information, with the effect-size contribution concentrated in the latter. Several mental health-related predictors were retained alongside age at comparably high frequency in at least one horizon, including psychotic-related prescribing, prior attended outpatient status, GP-coded schizophrenia, polypharmacy, substance dependence prescribing, hypnotic and anxiolytic prescribing, and alcoholic liver disease (each 80 to 100%). Non-demographic mental health-related features contributed reliably across resamples.

**Figure 4.**
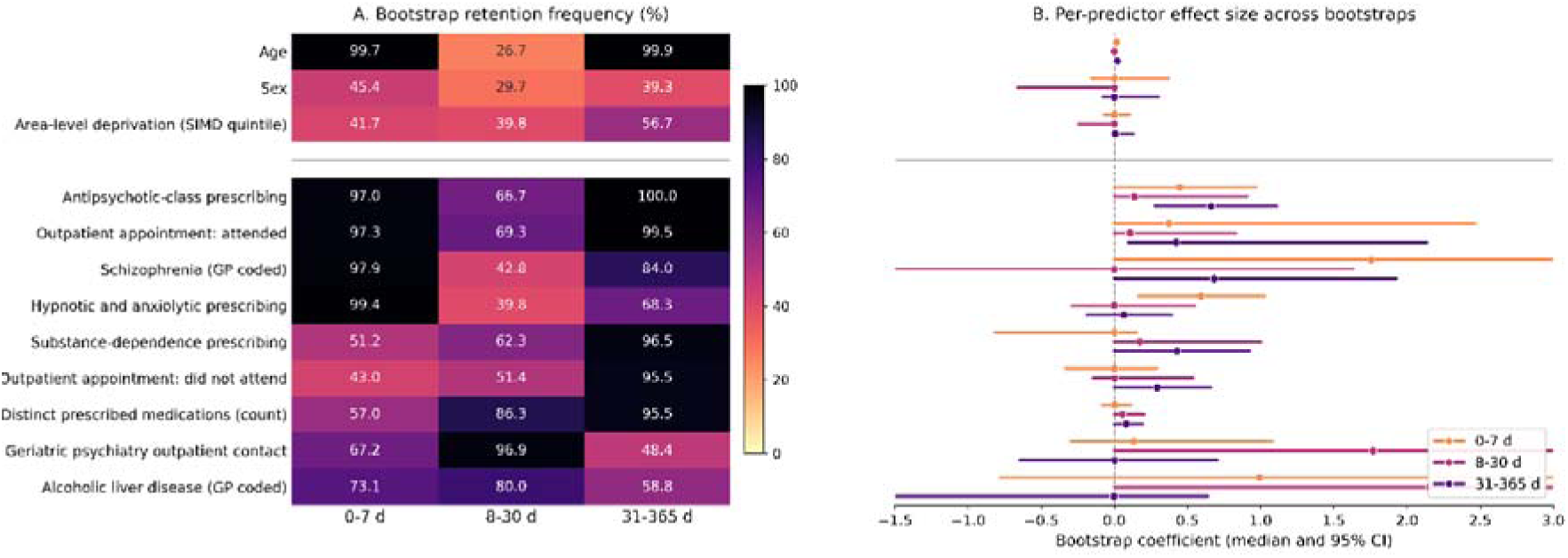
(A) Bootstrap retention frequencies (percent of 1,000 resamples of the training data) for demographic and mental health-related candidate predictors selected by the modelling approach, subset by follow-up window. (B) Bootstrap coefficient distributions (median and 95% confidence interval) for the same predictors on the elastic-net coefficient scale; note the disparity in effect size between age (retained at high frequency but with median coefficient near zero) and the mental health-related predictors. Intervals are clipped at the plot range; coefficients of exactly zero across all bootstraps are omitted. Complete retention tables for all 37 predictors are provided in the supplement.

## Discussion

### What this analysis shows

In this whole-population emergency department cohort of 4,095 adults presenting after paracetamol overdose, multivariable models built from routinely collected electronic health record data ranked patients above chance for the subsequent composite outcome of death or mental health admission, at every follow-up horizon examined. The bootstrap retention pattern reported above indicates that the signal driving this discrimination is not predominantly demographic.

Bootstrap-median calibration slopes were close to one and the 95% confidence interval on the calibration intercept included zero at every horizon, although with wide uncertainty at the lowest event rate. The same broad categories of feature (prior mental health service contact, psychiatric and substance dependence prescribing, polypharmacy, substance misuse markers) recurred across horizons in clinically face-valid configurations.

### What this analysis does not show

The models presented are not proposed for clinical use. The held-out discrimination of 0.70 to 0.86 across the three follow-up windows is below what would be required to credibly recognise an individual patient as having a higher risk of adverse outcome, and population-scale calibration of the kind reported here does not by itself license group-level intervention design. This work does not argue for replacing formulation-based psychosocial assessment: the recommended pathway addresses the psychological drivers of suicidal behaviour and the needs of the individual in ways that no model built on existing structured data can. We deliberately report discrimination and calibration only, and present no sensitivity, specificity or other threshold-based operating characteristics, precisely because we do not intend these models to be read as usable classifiers.

The position taken by current UK guidance is that risk assessment tools and stratification should not be used to predict suicide or repetition of self-harm, nor to determine who should be offered treatment or discharged; an emerging consensus extends this to the broader claim that prediction in this setting is not possible. Our results are inconsistent with that broader claim, and suggest there is potential risk signal which can be extracted from structured health data. We accept the empirical basis on which current guidance principally rests: at clinically plausible thresholds, prediction in this population yields low positive predictive value and a high false-positive burden, and stratification carries real risks of gate-keeping and false reassurance. Our results do not dispute this. The focussed point is that whether discriminative signal exists is a separate scientific question, and that current guidance about how tools should be used in practice should not be read as settling that question, or stopping ongoing enquiry which may change what is achievable by developing richer data and novel methods.

### Guidance trajectory and the risk of premature closure

The recommendation against using older risk assessment instruments is well-founded. The instruments audited by Graney and colleagues in 2020 were short, locally produced, mostly not derived from multivariable models, and used inconsistently across NHS organisations^11^. The base-rate critique by Pokorny and the 50-year meta-analysis by Franklin and colleagues addressed individual-level prediction from single risk factors^12,13^. None of those critiques addressed the discrimination achievable at population level from multivariable models built on linked routinely collected data.

The critique of currently available structured risk prediction tools is increasingly read as supporting a different and stronger claim: that no useful predictive signal exists in routinely collected data, and that prediction-based approaches as a class are unhelpful in self-harm. However, the evidence is conflicting.

Internationally, the same published evidence has supported different policy responses. In the United States, the Veterans Health Administration’s REACH VET predictive outreach has been scaled to all 140 facilities, though with known limitations including disparate performance by ethnicity and a low positive predictive value at the individual level^23–26^. Where the choice between responses is a policy judgement, the evidential inputs should remain open to revision rather than treated as settled. A categorical reading that “risk cannot be predicted” closes the underlying question at a moment when machine learning methods and new methods for extracting and integrating data are delivering meaningful change in other care settings.

Should sufficiently robust models emerge, translation of population-level signal into clinical use would raise implementation questions beyond predictive performance. Clinicians would need the data literacy to interpret probabilistic model outputs correctly, with consequent training and workforce implications. Safeguards would be needed so that model outputs broaden rather than gate-keep access to psychosocial assessment and intervention, and deployment would need to avoid the dynamics of blame documented around zero-suicide initiatives, in which staff and families are left feeling responsible for outcomes no assessment could have foreseen.

### Current routinely collected data may have a predictive ceiling

The level of discrimination we observed plausibly reflects an upper limit on what structured routinely collected EHR data of this kind can deliver, rather than a limitation of the modelling approach. The candidate feature set covered demographics, prior outpatient activity at specialty level, GP-coded diagnoses across the existing mental health phenotype list, and community prescribing at therapeutic moiety level. No major structured feature class was omitted. More flexible learners (gradient-boosted trees, random forests, neural networks) typically deliver modest gains at the cost of interpretability and, at this sample size, are unlikely to be transformative^27^. The question of how much additional signal is available is therefore predominantly a question about what new data could be added, not about which algorithm should be used.

The clinically interesting features are largely not in the structured data. Structured EHR records capture what was prescribed, attended and diagnosed; they do not capture the patient’s account of intent or precipitants, the clinician’s mental state findings, the specificity of any continuing ideation, the prominence of hopelessness, the patient’s described supports and protective factors, or the clinician’s gestalt judgement. These are precisely the variables that psychiatric assessment is designed to capture, and they are the features on which the existing recommended pathway, comprehensive psychosocial assessment by a mental health professional, actually relies. Each has factual support as a discriminator of subsequent outcome, and each is typically present in clinical free text without being available to any statistical model that consumes structured data alone.

Natural language processing of clinical free text is the most direct route to enlarging the structured feature space, and what it could expose corresponds closely to what the recommended psychosocial assessment pathway already treats as the substance of risk assessment^28^. Models that detect suicidal ideation from clinical or social media text reach 80 to 90% accuracy in controlled benchmarks^29^; signal recoverable from psychiatric clinical narratives has been shown to add discriminative performance over structured features alone^30^. A longer-term direction is the structured knowledge representation of clinical reasoning so that what the model learns from is not only the trace of what clinicians did but the constraints that bound their reasoning. The categorical reading of current English guidance, in its strongest form, would close off this category of work before it has been undertaken at scale, leaving patients with mental ill-health without the gains in predictive medicine being achieved in physical health domains^31,32^.

### Limitations

The cohort is drawn from a single NHS health board over a defined period, limiting generalisability to other healthcare settings. Re-presentations following an initial paracetamol overdose may have occurred outside this health board and would not be captured. Held-out test data are from the same period and geography as the training data; external validation in additional settings is not provided.

The composite outcome combines all-cause mortality and mental health admission; in the 0-7 day window in particular, this potentially includes deaths from the acute paracetamol toxicity itself, because follow-up is anchored to the index ED attendance rather than to discharge, and mode of death was not available for this study. The composite outcome was dominated by mental health admission at every horizon. Statistical disclosure control restrictions on small-count data (counts under 10) preclude release of the per-horizon breakdown of deaths versus mental health admissions; all component counts were non-zero. The discrimination reported here should therefore be read as predicting severe outcomes, death or deterioration requiring mental health admission, rather than as a direct prediction of suicide. The three horizons were modelled as separate prediction problems (reflecting how they would be prospectively applied by a clinician) rather than as a competing-risks or cumulative-incidence framework; this preserves the full at-risk denominator at each horizon but means the three horizon-specific models are not directly comparable to cumulative-incidence reports in the wider self-harm literature.

Because mental health admission dominates the composite and admission is itself a clinician-initiated decision, the models in part predict which patients the service subsequently acts on rather than an unmediated measure of deterioration, and low occurrence mortality-only outcomes cannot be reported separately under statistical disclosure control. We therefore frame the prediction target as the need for subsequent mental health intervention (a quantity the service already treats as actionable), and note that predicting it identifies the group toward which care is already directed.

We also note the analysis is observational and the models describe association between routinely recorded features and a composite adverse outcome rather than the effect of any intervention.

## Conclusion

Above-chance discriminative signal for adverse outcomes after paracetamol overdose was recoverable from routinely collected electronic health record data, in a single NHS health board cohort, at every horizon examined. Bootstrap retention indicated that this signal drew on mental health-related features alongside age, and not on age, sex and area-level deprivation alone. The emerging consensus that no useful predictive signal is recoverable from routinely collected data in this population is not borne out by the analysis presented here. The models themselves are not proposed for clinical use. Further work on richer data sources, particularly natural language processing of free-text clinical notes, is the route most likely to extend what was observed. Testing whether the same signal is present in all-method self-harm cohorts, and against suicide-specific outcomes where sample sizes allow, are the natural next steps. Whether suicide itself can be predicted from richer data remains unknown; it should be settled by evidence rather than closed by policy.

## Supporting information

Reporting checklist

## Declarations

### Contributors

Conceptualization, Methodology, Software: CH, JO; Validation: CH; Formal analysis: CH, JO; Investigation: CH, LS, JB; Resources: ME, AA; Data Curation: JO; Writing - original draft: CH, JO; Writing - review and editing: CH, LS, GB, JB, JO, AA, ME; Visualization, Supervision, Project administration: CH; Funding acquisition: CH, ME.

### Data availability statement

Individual-level data are not available outside the Scottish National Safe Haven secure research environment under DataLoch governance. Aggregate model outputs, calibration tables and bootstrap retention frequencies are provided in the supplement. Feature counts <10 are suppressed in line with DataLoch governance requirements. Analysis code is retained within the DataLoch trusted research environment and is not externally available.

### Ethics approval

DataLoch approval DL-2024-044.

