## Supplemental for "Reconsidering the case against risk prediction in self-harm: routinely collected health data distinguishes groups at higher and lower risk of adverse outcomes following paracetamol overdose"

**Supplementary Information**

*Reconsidering the case against risk prediction in self-harm: routinely collected health data can distinguish groups at differential risk across the cohort*

J Oxley, L Schölin, G Brennan, A Anand, J Brett, M Eddleston, C Humphries

### Contents

Table S1. Cohort flow.

Table S2. Training and held-out test set comparison, by 365-day composite outcome.

Table S3. Final hyperparameter values per model.

Table S4–S6. Full coefficient lists per model (retained predictors with coefficients and odds ratios).

Table S7–S9. Bootstrap retention frequencies and coefficient confidence intervals per model.

Figure S1–S3. Bootstrap feature retention frequency per model.

Figure S4–S6. Bootstrap coefficient dot-interval plots per model.

Table S10. Bootstrap retention frequencies for the full candidate predictor set, by follow-up window.

### Table S1. Cohort flow

*Numbers and changes through each filtering step from raw TrakCare emergency-attendance data to the final analysis cohort.*

| **Step** | **Attendances** | **Δ attendances** | **Patients** |
| --- | --- | --- | --- |
| Attendances with a matching discharge diagnosis code | 6,763 | - | 4,095 |
| Selecting the index (earliest) attendance per patient | 4,095 | -2,668 | 4,095 |

### Table S2. Training and held-out test set comparison

*Counts and proportions for each candidate feature, split by 80%/20% training/test sample and by 365-day composite outcome. P-values are from the corresponding chi-squared/Fisher (categorical) or Wilcoxon (continuous) tests on the training data and are provided for illustration only. "< 10" indicates a cell suppressed under NHS Scotland statistical disclosure control.*

| **Variable** | **Test, no event (n=714)** | **Test, event (n=106)** | **Training, no event (n=2,855)** | **Training, event (n=420)** |
| --- | --- | --- | --- | --- |
| Age (years) | 32 (23-48) | 44 (30.25-57) | 32 (24-47) | 41 (26.75-56) |
| GP diagnoses (count) | 0 (0-0) | 0 (0-1) | 0 (0-0) | 0 (0-1) |
| Polypharmacy (VTM count) | 1 (0-2) | 2 (1-5) | 1 (0-2) | 2 (1-4) |
| Sex: F | 434 (60.78%) | 66 (62.26%) | 1747 (61.19%) | 259 (61.67%) |
| Sex: M | 280 (39.22%) | 40 (37.74%) | 1108 (38.81%) | 161 (38.33%) |
| SIMD Quintile: 1 | 134 (18.77%) | 26 (24.53%) | 654 (22.91%) | 95 (22.62%) |
| SIMD Quintile: 2 | 221 (30.95%) | 44 (41.51%) | 833 (29.18%) | 109 (25.95%) |
| SIMD Quintile: 3 | 134 (18.77%) | 12 (11.32%) | 482 (16.88%) | 81 (19.29%) |
| SIMD Quintile: 4 | 101 (14.15%) | 14 (13.21%) | 396 (13.87%) | 62 (14.76%) |
| SIMD Quintile: 5 | 102 (14.29%) | 10 (9.43%) | 420 (14.71%) | 69 (16.43%) |
| SIMD Quintile: NA | 22 (3.08%) | NA | 70 (2.45%) | < 10 |
| SIMD: missing | 22 (3.08%) | < 10 | 70 (2.45%) | < 10 |
| Rx: Substance-dependence | 40 (5.6%) | 17 (16.04%) | 158 (5.53%) | 59 (14.05%) |
| Rx: Antiepileptics | 60 (8.4%) | 24 (22.64%) | 285 (9.98%) | 82 (19.52%) |
| Rx: Antidepressants | 335 (46.92%) | 70 (66.04%) | 1366 (47.85%) | 283 (67.38%) |
| Rx: Hypnotics / anxiolytics | 148 (20.73%) | 45 (42.45%) | 584 (20.46%) | 186 (44.29%) |
| Rx: Analgesics | 95 (13.31%) | 25 (23.58%) | 394 (13.8%) | 75 (17.86%) |
| Rx: Antipsychotic-class | 82 (11.48%) | 34 (32.08%) | 280 (9.81%) | 150 (35.71%) |
| Rx: CNS stimulant (ADHD) | < 10 | < 10 | 21 (0.74%) | < 10 |
| Any outpatient appt. | 297 (41.6%) | 75 (70.75%) | 1154 (40.42%) | 295 (70.24%) |
| Outpatient appt.: attended | 286 (40.06%) | 74 (69.81%) | 1103 (38.63%) | 291 (69.29%) |
| Outpatient appt.: not attended | 143 (20.03%) | 52 (49.06%) | 606 (21.23%) | 201 (47.86%) |
| Outpatient: General psychiatry | 275 (38.52%) | 70 (66.04%) | 1075 (37.65%) | 266 (63.33%) |
| Outpatient: Forensic psychiatry | < 10 | < 10 | 14 (0.49%) | < 10 |
| Outpatient: Geriatric psychiatry | 14 (1.96%) | < 10 | 34 (1.19%) | 29 (6.9%) |
| Outpatient: Clinical psychology | < 10 | < 10 | 34 (1.19%) | < 10 |
| Outpatient: Child & adolescent psychiatry | < 10 | < 10 | 37 (1.3%) | < 10 |
| Outpatient: Learning disability | < 10 | < 10 | 32 (1.12%) | < 10 |
| GP dx: Other psychiatric / substance misuse | 30 (4.2%) | < 10 | 122 (4.27%) | 28 (6.67%) |
| GP dx: Alcohol problems | 23 (3.22%) | < 10 | 118 (4.13%) | 44 (10.48%) |
| GP dx: Depression | 66 (9.24%) | 10 (9.43%) | 209 (7.32%) | 51 (12.14%) |
| GP dx: Anxiety | 50 (7%) | < 10 | 153 (5.36%) | 40 (9.52%) |
| GP dx: Personality disorder | 13 (1.82%) | < 10 | 71 (2.49%) | 33 (7.86%) |
| GP dx: Hyperkinetic disorder | < 10 | < 10 | < 10 | < 10 |
| GP dx: Anorexia / bulimia | < 10 | < 10 | < 10 | < 10 |
| GP dx: Dementia | < 10 | < 10 | < 10 | < 10 |
| GP dx: Schizophrenia | < 10 | < 10 | < 10 | 15 (3.57%) |
| GP dx: Autism spectrum | < 10 | < 10 | 13 (0.46%) | < 10 |
| GP dx: Delirium (non-substance) | < 10 | < 10 | < 10 | < 10 |
| GP dx: Alcoholic liver disease | < 10 | < 10 | < 10 | < 10 |
| GP dx: Intellectual disability | < 10 | < 10 | < 10 | < 10 |
| GP dx: Bipolar disorder | < 10 | < 10 | < 10 | < 10 |
| GP dx: OCD | < 10 | < 10 | < 10 | < 10 |

### Table S3. Final hyperparameter values

*Hyperparameter values selected by nested 5-fold cross-validation on the training set, maximising AUROC. α governs the mixing between LASSO and ridge regularisation (α=1 corresponds to pure LASSO); λ is the overall penalty strength.*

| **Window** | **α (mixing parameter)** | **λ (overall penalty)** | **Events / total (%)** |
| --- | --- | --- | --- |
| **0–7 days** | 0.8 | 0.0076 | 224 (5.47%) |
| **8–30 days** | 1.0 (pure LASSO) | 0.0027 | 82 (2.00%) |
| **31–365 days** | 0.5 | 0.0085 | 325 (7.94%) |

### Table S4. Full coefficient list, 0–7 day model

*All 37 candidate predictors with their elastic-net-fitted coefficient and corresponding odds ratio. Coefficients of exactly zero indicate predictors dropped by L1 regularisation.*

| **Feature** | **Coefficient (log-odds)** | **Odds ratio** |
| --- | --- | --- |
| (Intercept) | -3.233 | 0.04 |
| Age (years) | 0.002 | 1.00 |
| Sex | 0.000 | - |
| SIMD quintile | 0.000 | - |
| SIMD: missing | 0.000 | - |
| Polypharmacy (VTM count) | 0.041 | 1.04 |
| Rx: Substance-dependence | 0.000 | - |
| Rx: Antiepileptics | 0.000 | - |
| Rx: Antidepressants | 0.000 | - |
| Rx: Hypnotics / anxiolytics | 0.370 | 1.45 |
| Rx: Analgesics | 0.000 | - |
| Rx: Antipsychotic-class | 0.294 | 1.34 |
| Rx: CNS stimulant (ADHD) | 0.000 | - |
| Any outpatient appt. | 0.000 | - |
| Outpatient appt.: attended | 0.166 | 1.18 |
| Outpatient appt.: not attended | 0.000 | - |
| Outpatient: General psychiatry | 0.000 | - |
| Outpatient: Forensic psychiatry | 0.000 | - |
| Outpatient: Geriatric psychiatry | 0.000 | - |
| Outpatient: Clinical psychology | 0.000 | - |
| Outpatient: Child & adolescent psychiatry | 0.000 | - |
| Outpatient: Learning disability | 0.000 | - |
| GP diagnoses (count) | 0.000 | - |
| GP dx: Other psychiatric / substance misuse | 0.000 | - |
| GP dx: Alcohol problems | 0.000 | - |
| GP dx: Depression | 0.000 | - |
| GP dx: Anxiety | 0.000 | - |
| GP dx: Personality disorder | 0.000 | - |
| GP dx: Hyperkinetic disorder | 0.000 | - |
| GP dx: Anorexia / bulimia | 0.000 | - |
| GP dx: Dementia | 0.000 | - |
| GP dx: Schizophrenia | 1.105 | 3.02 |
| GP dx: Autism spectrum | 0.000 | - |
| GP dx: Delirium (non-substance) | 0.000 | - |
| GP dx: Alcoholic liver disease | 0.000 | - |
| GP dx: Intellectual disability | 0.000 | - |
| GP dx: Bipolar disorder | 0.000 | - |
| GP dx: OCD | 0.000 | - |

### Table S5. Full coefficient list, 8–30 day model

*All 37 candidate predictors with their elastic-net-fitted coefficient and corresponding odds ratio.*

| **Feature** | **Coefficient (log-odds)** | **Odds ratio** |
| --- | --- | --- |
| (Intercept) | -4.013 | 0.02 |
| Age (years) | 0.000 | - |
| Sex | 0.000 | - |
| SIMD quintile | 0.000 | - |
| SIMD: missing | 0.000 | - |
| Polypharmacy (VTM count) | 0.017 | 1.02 |
| Rx: Substance-dependence | 0.000 | - |
| Rx: Antiepileptics | 0.000 | - |
| Rx: Antidepressants | 0.000 | - |
| Rx: Hypnotics / anxiolytics | 0.000 | - |
| Rx: Analgesics | 0.000 | - |
| Rx: Antipsychotic-class | 0.000 | - |
| Rx: CNS stimulant (ADHD) | 0.000 | - |
| Any outpatient appt. | 0.000 | - |
| Outpatient appt.: attended | 0.000 | - |
| Outpatient appt.: not attended | 0.000 | - |
| Outpatient: General psychiatry | 0.000 | - |
| Outpatient: Forensic psychiatry | 0.000 | - |
| Outpatient: Geriatric psychiatry | 1.059 | 2.88 |
| Outpatient: Clinical psychology | 0.000 | - |
| Outpatient: Child & adolescent psychiatry | 0.000 | - |
| Outpatient: Learning disability | 0.000 | - |
| GP diagnoses (count) | 0.000 | - |
| GP dx: Other psychiatric / substance misuse | 0.000 | - |
| GP dx: Alcohol problems | 0.000 | - |
| GP dx: Depression | 0.000 | - |
| GP dx: Anxiety | 0.000 | - |
| GP dx: Personality disorder | 0.000 | - |
| GP dx: Hyperkinetic disorder | 0.000 | - |
| GP dx: Anorexia / bulimia | 0.000 | - |
| GP dx: Dementia | 0.000 | - |
| GP dx: Schizophrenia | 0.000 | - |
| GP dx: Autism spectrum | 0.000 | - |
| GP dx: Delirium (non-substance) | 0.000 | - |
| GP dx: Alcoholic liver disease | 0.429 | 1.54 |
| GP dx: Intellectual disability | 0.000 | - |
| GP dx: Bipolar disorder | 0.000 | - |
| GP dx: OCD | 0.000 | - |

### Table S6. Full coefficient list, 31–365 day model

*All 37 candidate predictors with their elastic-net-fitted coefficient and corresponding odds ratio.*

| **Feature** | **Coefficient (log-odds)** | **Odds ratio** |
| --- | --- | --- |
| (Intercept) | -3.097 | 0.05 |
| Age (years) | 0.004 | 1.00 |
| Sex | 0.000 | - |
| SIMD quintile | 0.000 | - |
| SIMD: missing | 0.000 | - |
| Polypharmacy (VTM count) | 0.098 | 1.10 |
| Rx: Substance-dependence | 0.190 | 1.21 |
| Rx: Antiepileptics | 0.000 | - |
| Rx: Antidepressants | 0.000 | - |
| Rx: Hypnotics / anxiolytics | 0.000 | - |
| Rx: Analgesics | 0.000 | - |
| Rx: Antipsychotic-class | 0.485 | 1.62 |
| Rx: CNS stimulant (ADHD) | 0.000 | - |
| Any outpatient appt. | 0.029 | 1.03 |
| Outpatient appt.: attended | 0.218 | 1.24 |
| Outpatient appt.: not attended | 0.263 | 1.30 |
| Outpatient: General psychiatry | 0.000 | - |
| Outpatient: Forensic psychiatry | 0.000 | - |
| Outpatient: Geriatric psychiatry | 0.000 | - |
| Outpatient: Clinical psychology | 0.000 | - |
| Outpatient: Child & adolescent psychiatry | 0.000 | - |
| Outpatient: Learning disability | 0.000 | - |
| GP diagnoses (count) | 0.002 | 1.00 |
| GP dx: Other psychiatric / substance misuse | 0.000 | - |
| GP dx: Alcohol problems | 0.000 | - |
| GP dx: Depression | 0.000 | - |
| GP dx: Anxiety | 0.000 | - |
| GP dx: Personality disorder | 0.000 | - |
| GP dx: Hyperkinetic disorder | 0.000 | - |
| GP dx: Anorexia / bulimia | 0.000 | - |
| GP dx: Dementia | 0.000 | - |
| GP dx: Schizophrenia | 0.000 | - |
| GP dx: Autism spectrum | 0.000 | - |
| GP dx: Delirium (non-substance) | 0.000 | - |
| GP dx: Alcoholic liver disease | 0.000 | - |
| GP dx: Intellectual disability | 0.000 | - |
| GP dx: Bipolar disorder | 0.000 | - |
| GP dx: OCD | 0.000 | - |

### Table S7. Bootstrap retention frequencies, 0–7 day model

*Across 1,000 bootstrap resamples of the training data, the proportion of iterations in which each candidate predictor was retained by the elastic-net penalty, alongside median coefficient and bootstrap 95% confidence interval. Sorted by retention frequency.*

| **Feature** | **Retention frequency (%)** | **Median coefficient** | **Lower 95% CI** | **Upper 95% CI** |
| --- | --- | --- | --- | --- |
| Age (years) | 99.7 | 0.015 | 0.003 | 0.028 |
| Rx: Hypnotics / anxiolytics | 99.4 | 0.592 | 0.163 | 1.028 |
| GP dx: Schizophrenia | 97.9 | 1.754 | 0.000 | 3.152 |
| Outpatient appt.: attended | 97.3 | 0.372 | 0.000 | 2.463 |
| Rx: Antipsychotic-class | 97.0 | 0.446 | 0.000 | 0.970 |
| GP dx: Depression | 73.1 | 0.224 | 0.000 | 0.811 |
| GP dx: Alcoholic liver disease | 73.1 | 0.991 | -0.780 | 3.129 |
| Rx: Antiepileptics | 71.1 | 0.127 | -0.050 | 0.617 |
| GP dx: Anorexia / bulimia | 69.7 | -0.421 | -5.315 | 0.000 |
| GP dx: Bipolar disorder | 69.5 | 0.346 | -1.374 | 2.037 |
| Outpatient: Geriatric psychiatry | 67.2 | 0.134 | -0.294 | 1.077 |
| Outpatient: Forensic psychiatry | 60.7 | 0.163 | -1.010 | 1.947 |
| GP dx: Autism spectrum | 57.7 | 0.000 | -4.796 | 0.497 |
| Polypharmacy (VTM count) | 57.0 | 0.002 | -0.083 | 0.115 |
| GP dx: Dementia | 55.6 | 0.000 | -2.277 | 1.652 |
| GP dx: Hyperkinetic disorder | 55.5 | 0.000 | -4.581 | 2.089 |
| Rx: Antidepressants | 51.9 | 0.000 | -0.107 | 0.449 |
| Rx: Analgesics | 51.4 | 0.000 | -0.653 | 0.032 |
| Rx: Substance-dependence | 51.2 | 0.000 | -0.819 | 0.151 |
| Outpatient: Clinical psychology | 50.5 | 0.000 | -1.235 | 1.119 |
| SIMD: missing | 49.6 | 0.000 | -4.175 | 0.000 |
| Outpatient: Child & adolescent psychiatry | 48.9 | 0.000 | -3.397 | 0.382 |
| Outpatient: Learning disability | 48.8 | 0.000 | -3.065 | 0.186 |
| GP dx: Personality disorder | 48.4 | 0.000 | -0.696 | 0.574 |
| Outpatient: General psychiatry | 47.2 | 0.000 | 0.000 | 0.845 |
| GP dx: Alcohol problems | 47.2 | 0.000 | -1.091 | 0.065 |
| Rx: CNS stimulant (ADHD) | 45.5 | 0.000 | -2.675 | 1.047 |
| Sex | 45.4 | 0.000 | -0.159 | 0.373 |
| GP dx: Delirium (non-substance) | 45.2 | 0.000 | -4.301 | 0.000 |
| Any outpatient appt. | 44.0 | 0.000 | -2.675 | 0.382 |
| GP dx: Anxiety | 43.5 | 0.000 | -0.434 | 0.558 |
| GP dx: Other psychiatric / substance misuse | 43.4 | 0.000 | -0.826 | 0.242 |
| Outpatient appt.: not attended | 43.0 | 0.000 | -0.335 | 0.292 |
| SIMD quintile | 41.7 | 0.000 | -0.071 | 0.103 |
| GP dx: Intellectual disability | 30.6 | 0.000 | -3.653 | 0.000 |
| GP diagnoses (count) | 16.0 | 0.000 | -0.028 | 0.104 |
| GP dx: OCD | 12.8 | 0.000 | -2.814 | 0.000 |

### Table S8. Bootstrap retention frequencies, 8–30 day model

*Across 1,000 bootstrap resamples of the training data, the proportion of iterations in which each candidate predictor was retained by the elastic-net penalty, alongside median coefficient and bootstrap 95% confidence interval. Sorted by retention frequency.*

| **Feature** | **Retention frequency (%)** | **Median coefficient** | **Lower 95% CI** | **Upper 95% CI** |
| --- | --- | --- | --- | --- |
| Outpatient: Geriatric psychiatry | 96.9 | 1.767 | 0.000 | 3.104 |
| Polypharmacy (VTM count) | 86.3 | 0.055 | 0.000 | 0.205 |
| GP dx: Alcoholic liver disease | 80.0 | 2.153 | 0.000 | 5.223 |
| GP dx: Anorexia / bulimia | 70.7 | 1.174 | -0.218 | 3.250 |
| Outpatient appt.: attended | 69.3 | 0.106 | 0.000 | 0.829 |
| Any outpatient appt. | 66.8 | 0.088 | 0.000 | 0.829 |
| Rx: Antipsychotic-class | 66.7 | 0.136 | 0.000 | 0.910 |
| GP dx: Autism spectrum | 63.1 | 0.756 | 0.000 | 3.032 |
| Rx: Substance-dependence | 62.3 | 0.174 | 0.000 | 1.003 |
| GP dx: Dementia | 61.7 | -0.339 | -3.043 | 0.000 |
| Outpatient appt.: not attended | 51.4 | 0.000 | -0.148 | 0.538 |
| GP dx: Alcohol problems | 47.7 | 0.000 | 0.000 | 1.277 |
| GP dx: Anxiety | 44.8 | 0.000 | 0.000 | 1.150 |
| GP dx: Schizophrenia | 42.8 | 0.000 | -1.977 | 1.630 |
| GP dx: Personality disorder | 41.7 | 0.000 | -0.250 | 0.960 |
| GP diagnoses (count) | 40.8 | 0.000 | 0.000 | 0.261 |
| SIMD quintile | 39.8 | 0.000 | -0.245 | 0.000 |
| Rx: Hypnotics / anxiolytics | 39.8 | 0.000 | -0.286 | 0.548 |
| Outpatient: Forensic psychiatry | 37.0 | 0.000 | -0.977 | 1.970 |
| GP dx: Other psychiatric / substance misuse | 35.3 | 0.000 | -0.336 | 0.920 |
| Outpatient: Clinical psychology | 34.3 | 0.000 | -1.476 | 1.133 |
| Rx: Antidepressants | 34.0 | 0.000 | -0.126 | 0.558 |
| Outpatient: General psychiatry | 32.4 | 0.000 | 0.000 | 0.612 |
| Outpatient: Child & adolescent psychiatry | 31.8 | 0.000 | -2.135 | 0.000 |
| Sex | 29.7 | 0.000 | -0.662 | 0.000 |
| Outpatient: Learning disability | 29.5 | 0.000 | -1.122 | 1.381 |
| Rx: Antiepileptics | 29.3 | 0.000 | -0.459 | 0.554 |
| GP dx: Delirium (non-substance) | 28.3 | 0.000 | -3.155 | 0.000 |
| GP dx: Bipolar disorder | 28.0 | 0.000 | -1.860 | 0.000 |
| Age (years) | 26.7 | 0.000 | -0.001 | 0.015 |
| Rx: Analgesics | 26.5 | 0.000 | -0.367 | 0.495 |
| Rx: CNS stimulant (ADHD) | 23.5 | 0.000 | -1.496 | 0.000 |
| GP dx: Depression | 21.3 | 0.000 | -0.697 | 0.194 |
| GP dx: Hyperkinetic disorder | 18.7 | 0.000 | -0.984 | 0.000 |
| SIMD: missing | 18.6 | 0.000 | -0.916 | 0.601 |
| GP dx: Intellectual disability | 8.3 | 0.000 | -0.935 | 0.000 |
| GP dx: OCD | 4.9 | 0.000 | -1.014 | 0.000 |

### Table S9. Bootstrap retention frequencies, 31–365 day model

*Across 1,000 bootstrap resamples of the training data, the proportion of iterations in which each candidate predictor was retained by the elastic-net penalty, alongside median coefficient and bootstrap 95% confidence interval. Sorted by retention frequency.*

| **Feature** | **Retention frequency (%)** | **Median coefficient** | **Lower 95% CI** | **Upper 95% CI** |
| --- | --- | --- | --- | --- |
| Rx: Antipsychotic-class | 100.0 | 0.660 | 0.275 | 1.111 |
| Age (years) | 99.9 | 0.018 | 0.006 | 0.033 |
| Outpatient appt.: attended | 99.5 | 0.421 | 0.094 | 2.136 |
| Rx: Substance-dependence | 96.5 | 0.428 | 0.000 | 0.925 |
| Polypharmacy (VTM count) | 95.5 | 0.080 | 0.000 | 0.195 |
| Outpatient appt.: not attended | 95.5 | 0.291 | 0.000 | 0.657 |
| GP dx: Alcohol problems | 91.5 | 0.465 | 0.000 | 1.021 |
| GP dx: Personality disorder | 87.4 | 0.397 | 0.000 | 1.140 |
| GP dx: Schizophrenia | 84.0 | 0.682 | 0.000 | 1.928 |
| GP dx: Anorexia / bulimia | 82.7 | 0.872 | 0.000 | 2.208 |
| GP dx: Delirium (non-substance) | 72.1 | -0.745 | -6.581 | 0.000 |
| Outpatient: Child & adolescent psychiatry | 70.6 | 0.647 | 0.000 | 1.973 |
| Rx: Hypnotics / anxiolytics | 68.3 | 0.064 | -0.185 | 0.390 |
| GP dx: Intellectual disability | 66.5 | 0.425 | -1.390 | 4.091 |
| SIMD: missing | 64.1 | -0.217 | -4.622 | 0.000 |
| GP dx: Alcoholic liver disease | 58.8 | -0.004 | -5.877 | 0.640 |
| SIMD quintile | 56.7 | 0.005 | -0.013 | 0.131 |
| Outpatient: General psychiatry | 56.7 | 0.028 | 0.000 | 0.848 |
| GP dx: Dementia | 55.3 | 0.000 | -1.431 | 1.699 |
| Rx: Antidepressants | 53.8 | 0.004 | -0.051 | 0.440 |
| GP dx: Bipolar disorder | 52.2 | 0.000 | -1.374 | 1.213 |
| Outpatient: Learning disability | 51.1 | 0.000 | -2.340 | 0.224 |
| Rx: CNS stimulant (ADHD) | 50.1 | 0.000 | -4.604 | 0.176 |
| GP dx: Autism spectrum | 50.1 | 0.000 | -2.063 | 1.240 |
| GP dx: Hyperkinetic disorder | 49.2 | 0.000 | -3.217 | 1.618 |
| Outpatient: Forensic psychiatry | 49.1 | 0.000 | -1.102 | 1.743 |
| Outpatient: Geriatric psychiatry | 48.4 | 0.000 | -0.649 | 0.704 |
| Outpatient: Clinical psychology | 48.4 | 0.000 | -0.729 | 0.829 |
| Rx: Antiepileptics | 45.8 | 0.000 | -0.323 | 0.322 |
| Any outpatient appt. | 45.0 | 0.000 | -2.385 | 0.254 |
| Rx: Analgesics | 40.9 | 0.000 | -0.552 | 0.041 |
| GP dx: Other psychiatric / substance misuse | 40.8 | 0.000 | -0.604 | 0.240 |
| Sex | 39.3 | 0.000 | -0.077 | 0.298 |
| GP diagnoses (count) | 39.0 | 0.000 | 0.000 | 0.180 |
| GP dx: Anxiety | 37.2 | 0.000 | -0.366 | 0.545 |
| GP dx: Depression | 35.0 | 0.000 | -0.520 | 0.134 |
| GP dx: OCD | 17.2 | 0.000 | -4.440 | 0.000 |

### Figure S1. Bootstrap feature retention frequency, 0–7 day model

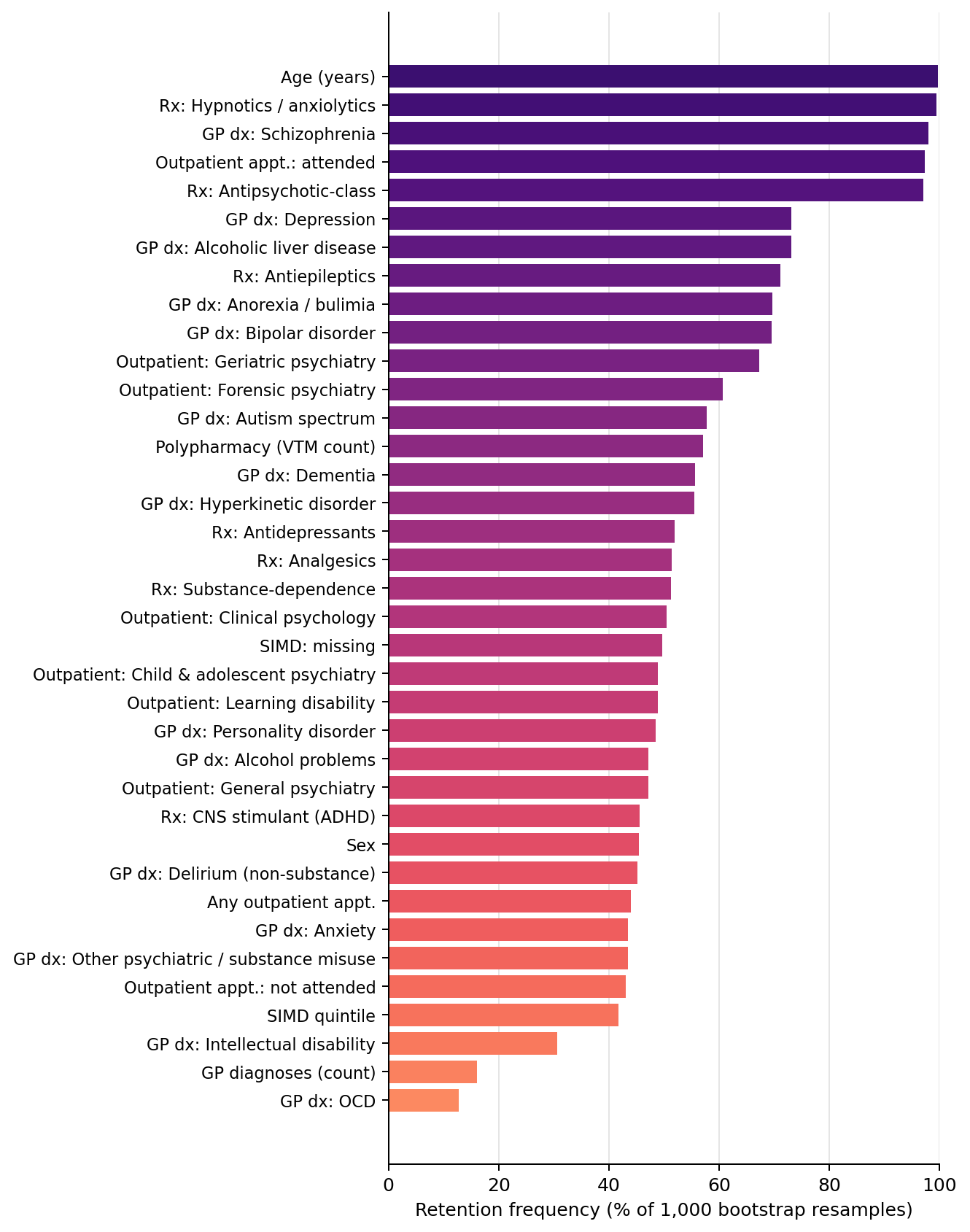

*Proportion of 1,000 bootstrap resamples of the training data in which each candidate predictor was retained by the elastic-net penalty (non-zero coefficient), 0–7 day model. Predictors are ordered by retention frequency.*

### Figure S2. Bootstrap feature retention frequency, 8–30 day model

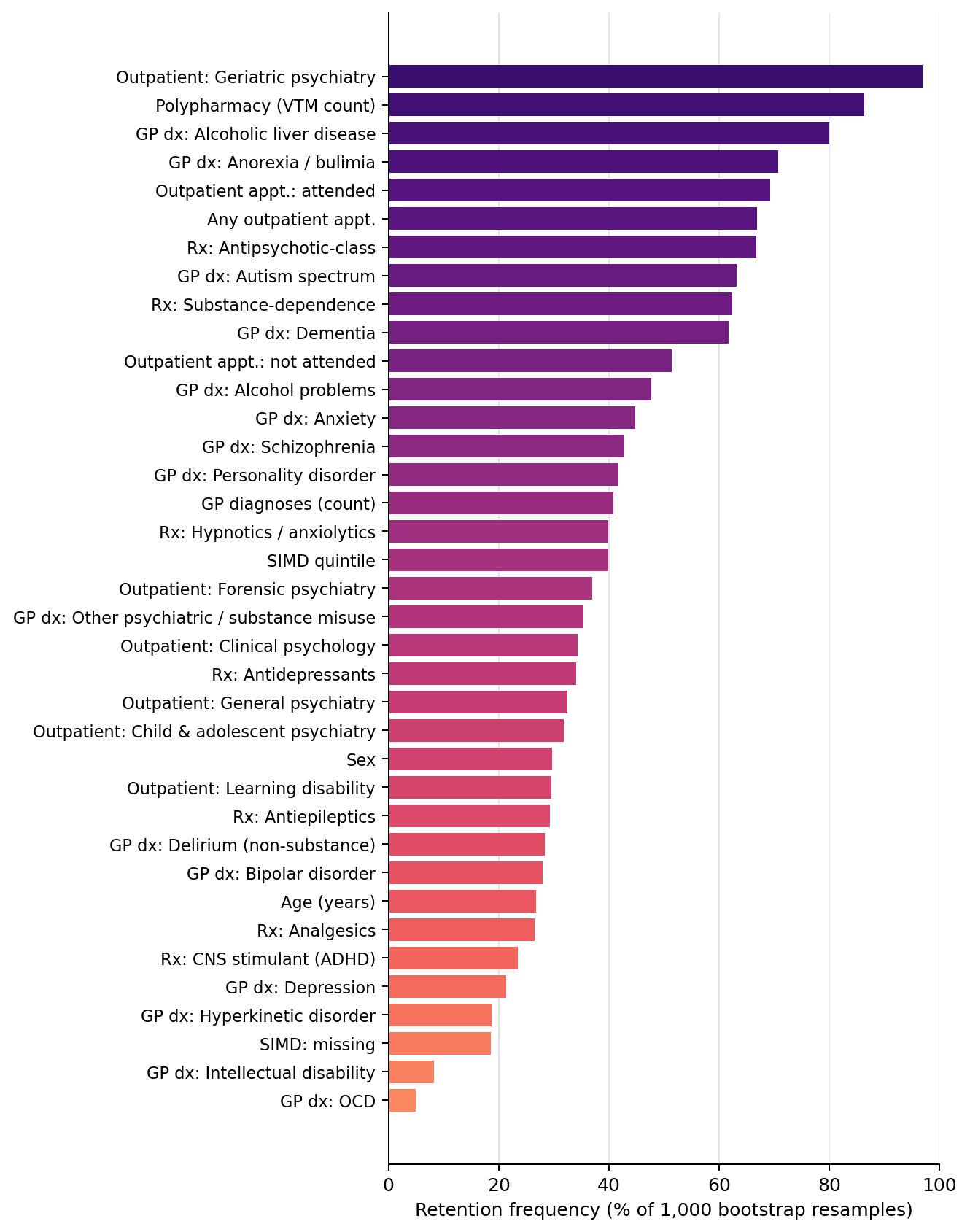

*Proportion of 1,000 bootstrap resamples of the training data in which each candidate predictor was retained by the elastic-net penalty (non-zero coefficient), 8–30 day model.*

### Figure S3. Bootstrap feature retention frequency, 31–365 day model

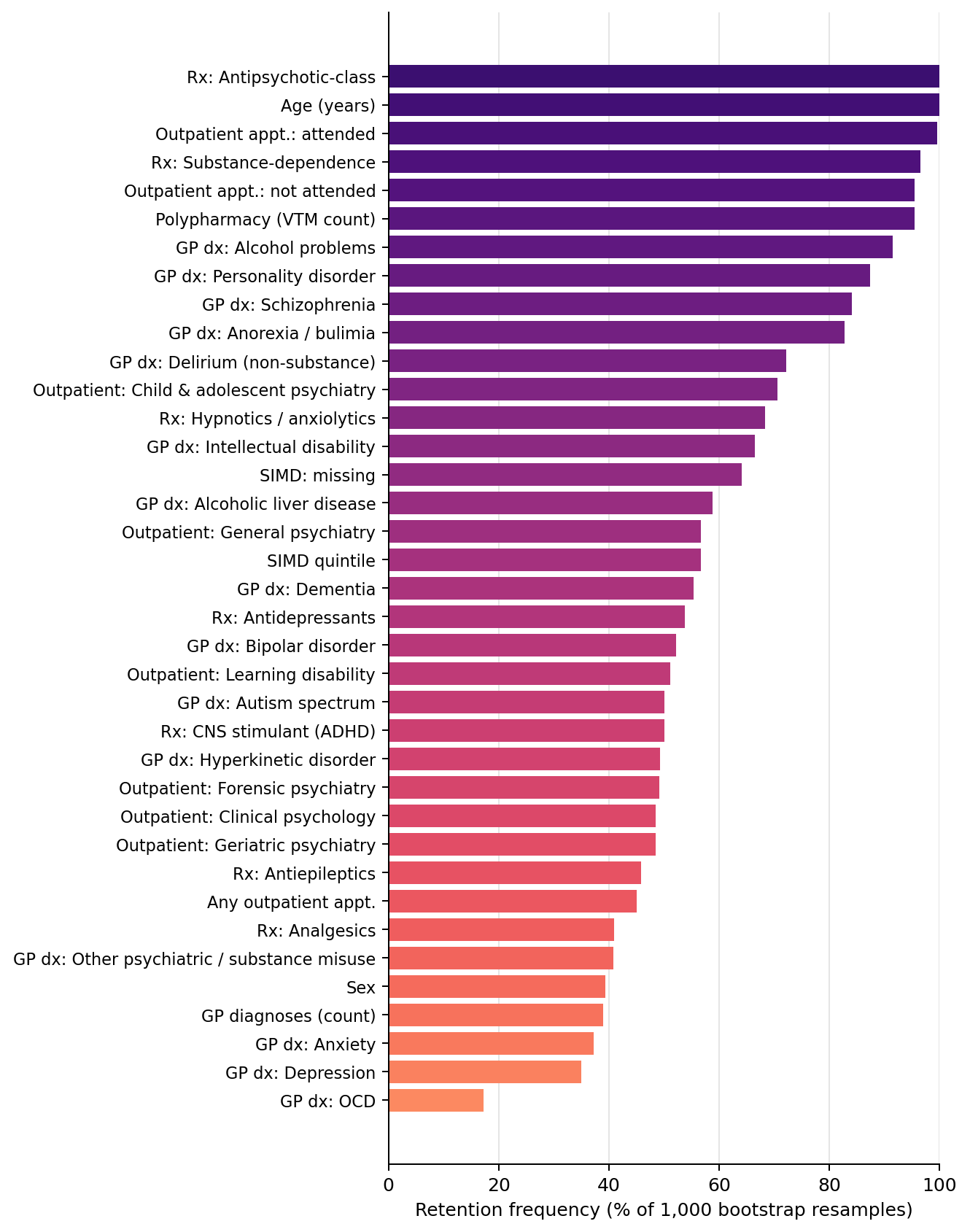

*Proportion of 1,000 bootstrap resamples of the training data in which each candidate predictor was retained by the elastic-net penalty (non-zero coefficient), 31–365 day model.*

### Figure S4. Bootstrap coefficient dot-interval plot, 0–7 day model

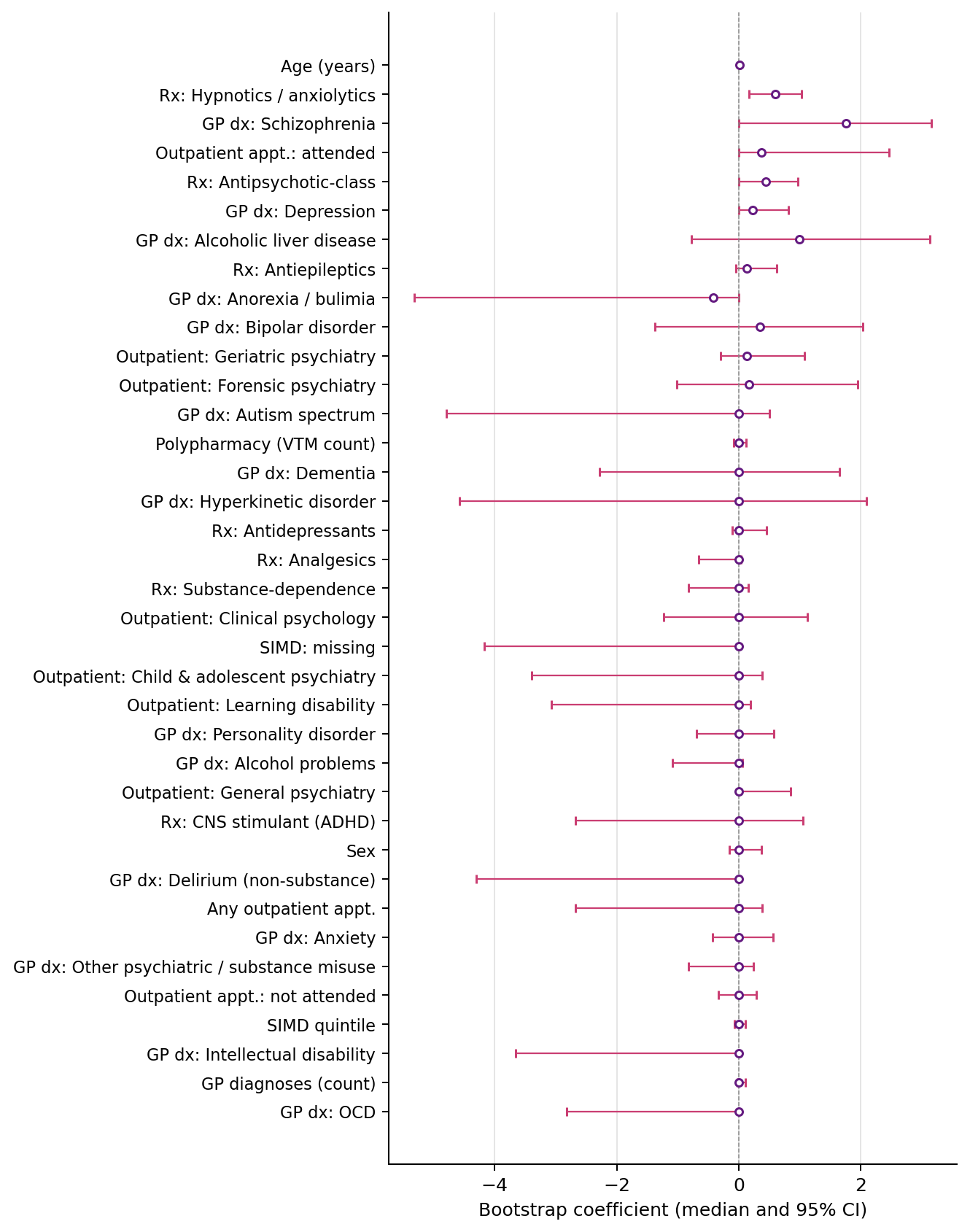

*Median and 95% confidence interval of the coefficient for each candidate predictor across 1,000 bootstrap resamples of the training data, 0–7 day model. Predictors ordered by retention frequency.*

### Figure S5. Bootstrap coefficient dot-interval plot, 8–30 day model

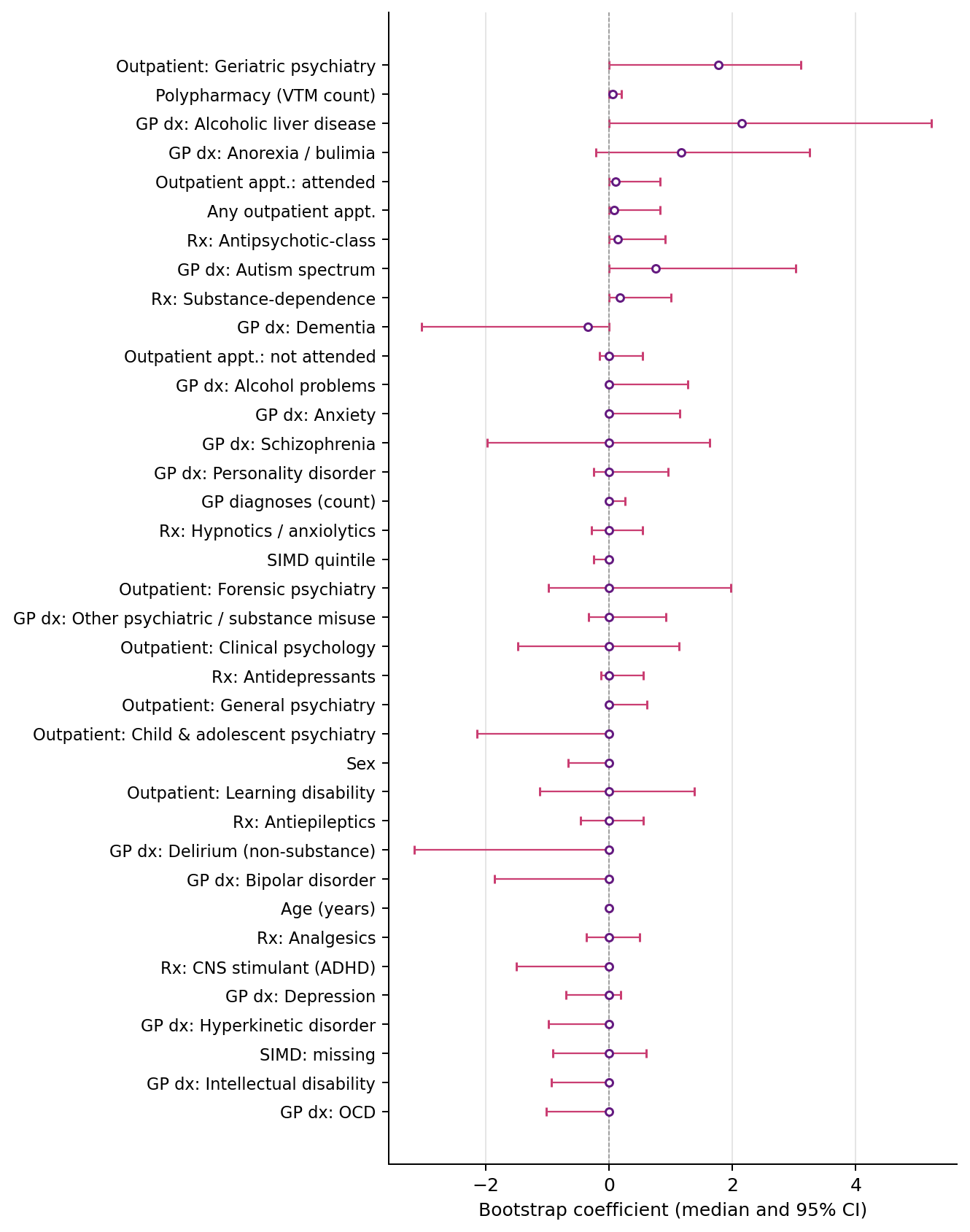

*Median and 95% confidence interval of the coefficient for each candidate predictor across 1,000 bootstrap resamples of the training data, 8–30 day model.*

### Figure S6. Bootstrap coefficient dot-interval plot, 31–365 day model

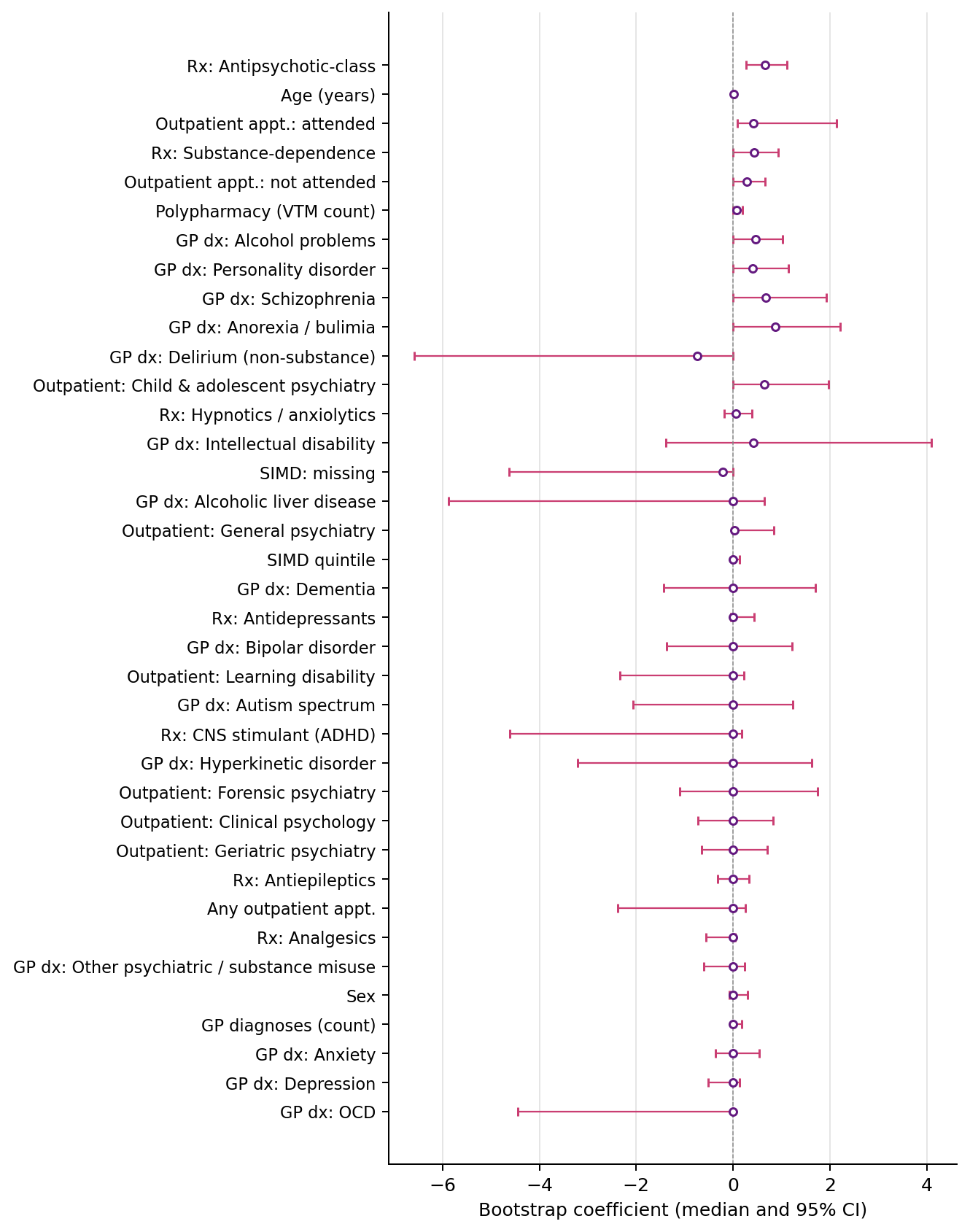

*Median and 95% confidence interval of the coefficient for each candidate predictor across 1,000 bootstrap resamples of the training data, 31–365 day model.*

### Table S10. Categories of feature retained per follow-up window

*Illustrative classification of retained features (non-zero coefficients) from the final elastic-net models. Originally Table 3 of the main paper. All retained coefficients carried positive sign (associated with increased odds of the composite outcome). Specific features should be interpreted as illustrative of the feature classes the model draws on, not as a definitive ranking.*

| **Feature category (illustrative)** | **0–7 days** | **8–30 days** | **31–365 days** |
| --- | --- | --- | --- |
| Age | retained (+) | - | retained (+) |
| Polypharmacy (unique VTM count) | retained (+) | retained (+) | retained (+) |
| Prior MH-specialty outpatient contact | retained (+) | retained (+) | retained (+) |
| Antipsychotic prescribing | retained (+) | - | retained (+) |
| Hypnotic / anxiolytic prescribing | retained (+) | - | - |
| Substance-dependence prescribing | - | - | retained (+) |
| Severe MH GP diagnosis (schizophrenia) | retained (+) | - | - |
| Physical multimorbidity (alcoholic liver disease) | - | retained (+) | - |
| Total GP diagnoses (multimorbidity) | - | - | retained (+) |
