## Supplementary material for "Reconsidering the case against risk prediction in self-harm: routinely collected health data distinguishes groups at higher and lower risk of adverse outcomes following paracetamol overdose": Reporting checklist

### TRIPOD+AI checklist

| **Item** | **Section** | **Checklist item** | **Reported on page / section** |
| --- | --- | --- | --- |
| 1 | Title | Identify the study as developing or evaluating multivariable prediction model(s). | Title |
| 2 | Abstract | Provide a structured summary (objectives, design, setting, participants, outcomes, methods, results, conclusions). | Abstract, p.1 |
| 3a | Background | Explain the medical context and rationale for developing/validating the model. | Introduction |
| 3b | Objectives | Specify the study objectives, including whether the study describes the development or validation of the model, or both. | Introduction (final paragraph) |
| 4a | Source of data | Describe the source of data, and basic details of the study design. | Methods: Cohort and data sources |
| 4b | Source of data | Specify the key dates of data collection. | Methods: Cohort (2017–2023) |
| 5a | Participants | Specify key elements of the setting (e.g., secondary care). | Methods: Cohort |
| 5b | Participants | Describe eligibility criteria for participants. | Methods: Cohort |
| 6a | Outcome | Clearly define the outcome that is predicted, including how and when assessed. | Methods: Outcome definition |
| 7a | Predictors | Clearly define all predictors used in developing the model. | Methods: Candidate predictor set; full list in Supplement S1 |
| 8 | Sample size | Explain how the study sample size was arrived at. | Methods: Sample-size considerations |
| 9 | Missing data | Describe how missing data were handled. | Methods: Missing-data handling |
| 10a | Statistical analysis | Describe how predictors were handled in the analysis. | Methods: Modelling approach |
| 10b | Statistical analysis | Specify type of model, modelling-building procedures, and method for internal validation. | Methods: Modelling approach + Bootstrap procedure |
| 10c | Statistical analysis | Specify any model performance measures used. | Methods: Performance evaluation |
| 12 | Risk groups | If risk groups defined, specify how they were derived. | N/A - no clinical risk groups derived |
| 13a | Participants | Describe the flow of participants through the study (including a flow diagram). | Results: cohort flow in Supplement Table S1 |
| 14a | Model development | Specify number of participants and outcomes for analysis. | Results: Cohort characteristics |
| 15a | Model specification | Present the final prediction model. | Results: Figure 3 retained coefficients |
| 16 | Model performance | Report performance measures (including calibration and discrimination). | Results: Figures 1 (ROC), 2 (calibration) |
| 17 | Model-updating | Report results of any updates of the model. | N/A |
| 18 | Limitations | Discuss any limitations of the study. | Discussion: Limitations |
| 19 | Interpretation | Give an overall interpretation of the results, considering objectives, limitations, results from similar studies, and other relevant evidence. | Discussion: Interpretation |
| 20 | Implications | Discuss the potential clinical use of the model and implications for future research. | Discussion: Implications; Conclusion (no clinical-use proposal) |
| 21 | Supplementary information | Provide information about the availability of supplementary resources. | Data sharing statement |
| 22 | Funding | Give the source of funding and the role of the funders for the present study. | Funding |
| TRIPOD-AI 1 | Fairness | Describe assessment of model performance across patient subgroups. | Discussion: Limitations |
| TRIPOD-AI 2 | Open science | Code, model artefacts and trained-model availability. | Data sharing statement |
| TRIPOD-AI 3 | Reporting bias | Address risk-of-bias considerations (PROBAST). | Discussion: Limitations |
